# Health-Related Quality of Life in Adults with Congenital Heart Disease - A Population-Based Study from the Australian National Registry

**DOI:** 10.1101/2025.10.17.25338261

**Authors:** Tanya Badal, William Baxter, Larissa Lloyd, Sasha Ruban, Calum Nicholson, Geoff Strange, Claudia Rutherford, David S. Celermajer

**Affiliations:** University of Sydney, Camperdown, NSW, 2050, Australia; Heart Research Institute, 7 Eliza St, Newtown NSW 2042, Australia; Royal Prince Alfred Hospital, 50 Missenden Rd, Camperdown NSW 2050, Australia

**Keywords:** heart defects, congenital, Patient Reported Outcome Measures, Quality of Life

## Abstract

**Background:** As survival into adulthood improves for congenital heart disease (CHD) patients, the quality of survival is paramount. We assessed health-related quality of life (HRQL) in a large Australian adult CHD cohort and identified clinical, psychosocial and system-level predictors of better or worse HRQL.

**Methods:** We conducted a multicenter, registry-based cross-sectional study of a random stratified sample of adults from the Registry, assessing 868 participants who completed the PedsQL 4.0 Generic Core Scales and had complete covariate data. Analyses were structured into three stages: Stage I examined CHD complexity (mild, moderate, severe) across the lower (bottom 15%), middle (70%), and upper (top 15%) HRQL score bands; Stage II identified predictors of worse or better HRQL at the tails of the distribution; and Stage III assessed predictors of overall HRQL within the severe CHD group.

**Results:** PedsQL Total scores were generally high (median 76.1, IQR 64.1–87.0) and best in Social functioning; Emotional and Psychosocial scales showed longer lower tails. Severe CHD was over-represented in the lower 15% and under-represented in the upper 15% bands (Total HRQL: *p* < 0.001; Cramér^′^s V = 0.10, 95% CI [0.06, 0.15]). At the 15th percentile, worse HRQL was associated with severe CHD (≈ −5.9 points), poor transition support (≈ −11.2), mood disorder (≈ −12.7), and diabetes (≈ −16.6). At the 85th percentile, severe CHD (≈ −5.2) and mental-health comorbidity (mood ≈ −7.3; anxiety ≈ −5.6) was associated with worse HRQL. In severe CHD, poor transition support and mood or anxiety disorders (*OR* ≈ 0.48, 0.30, and 0.36, respectively; p ≤ 0.01) markedly increased the odds of worse HRQL and reduced the odds of better HRQL. Very well-supported transition (*OR* ≈ 1.91, posterior probability = 0.99) was associated with better HRQL.

**Conclusion:** In adult CHD, anatomic complexity contributes modestly to HRQL extremes. Modifiable predictors (transition experience and mental-health comorbidity, location and diabetes) emerge as key targets to improve HRQL, particularly in severe CHD.

**What is Known; What the Study Adds:** *What is Known:*

- As survival into adulthood increases for congenital heart disease (CHD) patients, health-related quality of life (HRQL) is an important outcome, yet adult-focused evidence remains limited.
- Prior studies rely on mean-based analyses that obscure patients doing particularly poorly or well and are confounded by healthcare-system differences.

*What the Study Adds:*

- This is the first study in adult congenital heart disease (ACHD) research to use distribution-aware statistics (quantile regression and Bayesian proportional-odds models) to assess health-related quality of life (HRQL) beyond average scores and simple cutoffs to highlight risks and protective factors that may otherwise be missed.
- Poor transition support, mental-health comorbidity, diabetes, and regional or remote residence were key drivers of very low HRQL beyond anatomical severity, indicating ACHD-specific needs rather than disparities in healthcare access within Australia’s universal Medicare system.
- Clinical take-home: prioritize structured transition, integrated mental-health care, diabetes management, and equitable access for regional/remote patients, using distribution-aware targeting to identify those at greatest risk of worse HRQL.

## Introduction

The global incidence of congenital heart disease (CHD) over the recent decade is reported as 9 per 1,000 live births, with CHD contributing 6.4% to the total disease burden among infants^1^. Despite this, advances in surgical techniques, medical technologies and pediatric cardiac care have appreciably reduced mortality in children with CHD^2,3^,with now approximately 95% survival rates into adult life. Consequently, the global adult CHD (ACHD) population is steadily increasing, with survival rates improving and comorbidities accumulating with transitions into adulthood^4–8^. This shift is also evident within the Australian context, with an annual reported increase of approximately 5% in the ACHD population^9^. Considering this, research efforts have since focused on characterizing the association between CHD and lifelong comorbidities, and the impact on healthcare utilization^5^. Particularly, growing emphasis has been placed on evaluating the overall quality of survival^10^.

From a disease standpoint, health-related quality of life (HRQL) has become a core patient-centered outcome. Complementary to conventional biomedical metrics, HRQL captures domains specific to lived experience, including functioning, psychosocial wellbeing, and symptom burden^11^. A range of measures have been used to quantify and evaluate HRQL in CHD^11,12^. Among these, the PedsQL family of measures has consistently demonstrated strong psychometric properties across pediatric and adolescent populations^13^. More recently, its application has been extended through the development of health-state classification systems that enable preference-based valuation^14^.

Despite growing recognition of HRQL as a key outcome, ACHD literature has several limitations. Many earlier studies were constrained by small, single-center cohorts, restricting statistical power and limiting the ability to generalize findings nationally or internationally^10^. Such designs also rarely incorporated broader contextual factors, even though cross-country comparisons have shown that health-system and socioeconomic conditions substantially influence HRQL^11,12^. In addition, while domain-specific analyses consistently highlight cognitive and educational challenges in CHD, these patterns can be less visible when only global indices are reported^15,16^. This underscores that domain-specific and global approaches provide complementary insights. Finally, interpretation of pooled evidence is complicated by methodological heterogeneity, including differences in measures used, age ranges, and comparison groups as well as publication bias, whereby studies showing significant deficits are more likely to be published^15,16^.

In light of this, several large-scale studies have attempted to address these limitations by refining their analytical approaches. For instance, international multi-center surveys of ACHD patients found that overall HRQL in adulthood is generally good, with variation in HRQL largely attributable to individual patient characteristics rather than country-level factors^12,17^. Likewise, a new patient-driven registry of over 4,500 U.S. adults with CHD reported that 84% of participants rate their life quality as good or better regardless of defect complexity, while 88% live with one or more comorbid conditions (e.g. arrhythmias in one-third, mood disorders in over one-third)^18^. These contemporary findings illuminate both the improved outlook for many ACHD survivors and the persistent burden of health issues in a subset. However, the analytical methods in these studies–and those prior–have emphasized mean scores or broad group comparisons, which risk obscuring important within-group variability. To more effectively capture nuances at the extremes of the HRQL spectrum, investigators have started to apply more advanced distributional modelling. Most recently, quantile regression (QR) was used to identify predictors associated with particularly worse versus better HRQL by examining outcome percentiles rather than mere averages^19^. Similarly, cardiovascular research has utilized Bayesian proportional-odds (PO) models to compare patient-reported outcomes (PROs) across their entire ordinal distribution, using full posterior inference. This approach extends past a single average cutoff, yielding a more granular understanding of outcome differences across populations^20^. By leveraging such advanced approaches, researchers can more rigorously discern those predictors of especially better or excellent HRQL that otherwise can be masked by analyses employing classic comparisons of mean scores or simple regressions.

The present study overcomes these limitations and technical shortcomings by investigating HRQL in a large national cohort of Australian adults with CHD using the PedsQL 4.0 Generic Core Scales^21^ (GCS) Young Adult and Adult forms. Unlike most prior work, we explicitly examine the distributional tails of HRQL scores, identifying those at the lowest and highest extremes. Through a three-stage analytic framework combining banding, quantile regression, and Bayesian proportional-odds modelling, we assess not only defect severity but also psychosocial and system-level HRQL predictors associated with HRQL, including transition support, comorbidities, and location. To our knowledge, this is the first study in ACHD patients to employ such advanced distributional modelling in the evaluation of HRQL outcomes. Our approach improves upon previous research by avoiding reliance on mean-based regression or dichotomization, enabling a distribution-sensitive analysis that reveals how these predictors shift and influence the likelihood of belonging to the lowest or highest HRQL bands. In doing so, this work provides a more nuanced understanding of who fares poorly or well in adulthood with CHD and highlights modifiable predictors most relevant to clinical care and health policy.

## Methods

### Study Population

This cross-sectional prospective study employed a stratified random sample of patients^22^ from the Australian National CHD Registry, a multicenter database of over 68,000 CHD patients of all ages, across 10 tertiary centers^23^. The stratified sample was used to establish a profiling cohort (n=2,046, Supplemental Material Figure S1) to characterize the whole-of-life burden of CHD^22^. Patients were recruited from two adult and two pediatric centers, stratified by CHD complexity, age, sex, and location (urban vs. regional/remote). Participants gave written consent and completed multiple physical, neurocognitive, and patient-reported assessments, including the PedsQL GCS analyzed in this study. Surveys were administered through REDCap^24,25^.

Human Research Ethics Approval was obtained from the Sydney Local Health District Ethics Review Committee (RPAH Zone, EC00113) under protocol number 2020/ETH02832. The study adheres to the STROBE guidelines for the reporting of cohort data ^26^. The authors will not make study data openly available. Any future use of de-identified data will require specific ethics and governance approvals and will be limited to approved collaborators.

From the profiling cohort, 868 adults (≥ 18 years) completed the PedsQL GCS (adult or young adult versions) and had complete sociodemographic and comorbidity information; these individuals comprised the analytic cohort (Table 1). The median age was 39.7 years, with balanced sex distribution. CHD complexity was classified as 23% mild, 52% moderate, and 25% severe. While most participants resided in metropolitan areas, 25% were from regional/remote locations. Additional predictors were prespecified and included sociodemographic factors (marital status, dependents, travelling further than 200km to cardiology appointments, perception of transition support from pediatric to adult cardiac care) and comorbidities (diagnosed mood or anxiety disorder, diabetes, any form of cancer).

**Table 1:**
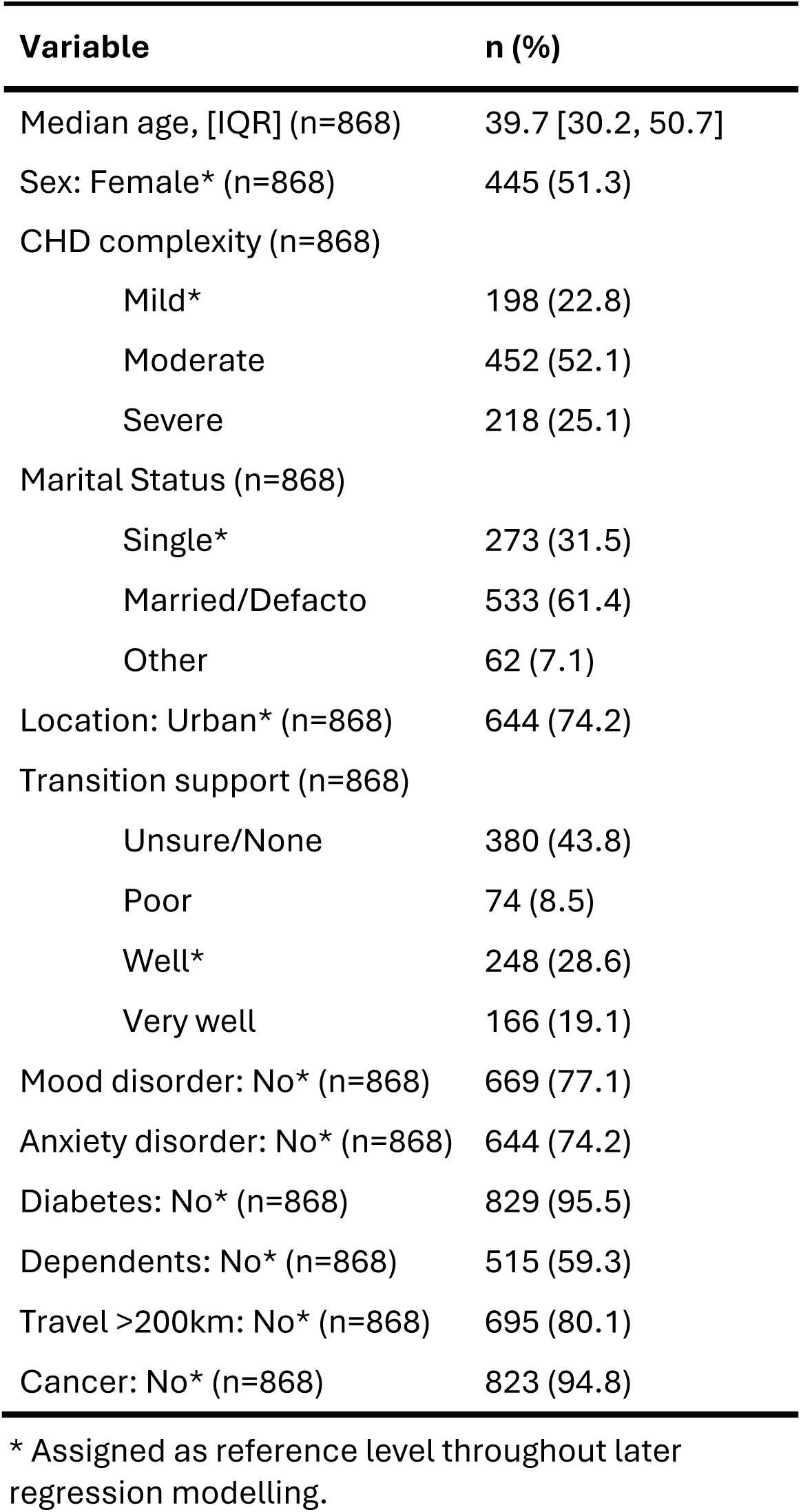
Sociodemographic and comorbidity variables for the total sample of adults with congenital heart disease (n=868).

### Health-Related Quality of Life Measure

We used the Young Adult (18–25 years) and Adult (≥ 26 years) versions of the PedsQL GCS, both comprising 23 items across four domains: Physical (8 items), Emotional (5), Social (5), and Work (5; adapted from “School” in the pediatric forms). All measure items reference the past month and are scored on a 5-point Likert scale (0: ‘Never’, 1: ‘Almost never’, 2: ‘Sometimes’, 3: ‘Often’, 4: ‘Almost always’). In accordance with the published PedsQL GCS scoring instructions, responses were reverse-scored and linearly transformed to a 0–100 scale, where higher values indicate better HRQL. Scale scores were calculated as the mean of available items provided, more than half were completed. In addition to four domain scales, we derived a Psychosocial Health summary scale score (Emotional, Social, Work) and a Total scale score (all domains combined).

### Analytic Framework

The analysis plan was determined prospectively to answer clinically relevant questions about HRQL – to assess the overall results; and to assess the predictors of particularly better or worse HRQL. Thus, we focused our analysis on the extremes of the distributions. Analyses were conducted in three stages:

- **Stage I:** Representation of CHD complexity across HRQL bands.
- **Stage II:** Predictors of overall HRQL at the lower and upper bands.
- **Stage III:** Predictors of overall HRQL upper-band membership within the severe CHD group.

For Stages II and III, the primary outcome was restricted to the PedsQL Total score. Stage III was further restricted to patients with severe CHD (n=218), given their higher risk of complications and poorer outcomes. Cut-points defining the lower and upper bands of each HRQL domain were respectively demarcated by the 15th and 85th percentiles (approximating ±1 standard deviation (SD) under normality), thereby capturing patients doing particularly better or worse.

#### Stage I: Representation of CHD complexity across HRQL bands

For each domain, patients were assigned to lower (bottom 15%), middle (middle 70%), or upper (top 15%) HRQL bands. We then compared the distribution of mild, moderate, and severe CHD across them using Pearson *χ*^2^ tests, determining whether severe CHD patients were more likely to fall in the lower band and mild CHD patients in the upper band. We report the resulting *χ*^2^ test statistics, with Cramér’s V and 95% confidence intervals (CIs) used to quantify effect size measures of association.

#### Stage II: Predictors of overall HRQL at the lower and upper bands

To investigate which variables were associated with worse or better HRQL scores, we fitted quantile regression (QR) models at the 15th and 85th percentiles of the PedsQL Total score. This approach allowed us to estimate how predictors influenced outcomes among patients with the best or worse reported HRQL, rather than only the mean. Predictors included the prespecified sociodemographic and clinical variables listed in Table 1, with reference categories as indicated. Age was modelled as a continuous variable and scaled per 10 years. Model coefficients are reported as shifts in Total score units at each percentile, with 95% CIs obtained using non-parametric bootstrap resampling. Assumptions of linearity and model adequacy were checked, and sensitivity analyses confirmed robustness.

#### Stage III: Predictors of overall HRQL upper-band membership within the severe CHD group

Within the severe CHD subgroup, we used a Bayesian cumulative-logit proportional-odds (PO) regression model to assess how patient and clinical factors influenced the odds of belonging to the lower, middle, or upper bands of the PedsQL Total Scale. Predictors were the same as in Stage II, with age excluded due to non-significance in those models. As severe CHD is both clinically central and relatively sparse in the tails, we partially pooled across CHD complexity strata by including random intercepts and slopes by complexity; thus the severe specific effect equals the population (fixed) effect plus the severity-level deviation. To mitigate the imbalance across bands, we used inverse-frequency case weights (normalized to a mean of 1). Partial pooling was used to control for confounding due to constrained sample sizes across CHD complexity. Results are expressed as odds ratios with 95% credible intervals (CrIs), where values >1 indicate greater odds of being in a higher HRQL band. PO assumptions were examined using a category-specific model, and model fit was confirmed by cross-validation.

### Software and Data Analysis

All data were collected and managed using REDCap^24,25^ electronic data capture tools hosted at the Sydney Local Health District (v 14.5.21) and analyses were performed in R ^27^(v4.3) using RStudio. Data wrangling and graphics used tidyverse^28^ (v2.0.0) and ggplot2^29^ (v3.5.2). Stage I used Pearson’s χ² tests with Cramér’s V as the effect size and bootstrap 95% CIs. Stage II used quantile regression via quantreg (v6.1), modelling age with natural cubic splines (splines v4.4.3); uncertainty was summarized with bootstrap 95% CIs. Stage III used a cumulative-logit proportional-odds model fitted in a Bayesian framework with brms (v2.22.0); results are reported as posterior medians with 95% credible intervals, with proportional-odds checks via a category-specific alternative and Pareto Smoothed Importance Sampling Leave-One-Out (PSIS-LOO) model comparison. Complete-case analysis was applied; two-sided tests used *α* = 0.05, and reporting emphasized effect sizes (Cramér’s V) and interval estimates. Full priors, sampler settings, diagnostics, and sensitivity analyses are provided in the Supplementary Material; analysis scripts are available to qualified investigators subject to registry governance. Finally, one author had full access to the study data and takes responsibility for its integrity and the data analysis.

## Results

### Score Distributions

Across the cohort (n=868), score distributions for all PedsQL GCS scales were generally high and skewed toward the upper range (Figure 1). Median (IQR) scores were 89.1 (79.7– 95.3) for Physical, 76.0 (64.0–88.0) for Emotional, 88.0 (76.0–100.0) for Social, 80.0 (68.0–92.0) for Work, 76.7 (65.0–88.3) for Psychosocial, and 76.1 (64.1–87.0) for the Total scale. The density gradient (yellow indicating low, purple indicating high) showed most patient’s scores concentrated in the upper range for the Physical and Social scales, with broader, lower-centered distributions for Emotional, Psychosocial and Total. The 15th/85th percentile cut-points bordering the lower/upper HRQL score bands were approximately 75/98.4 for Physical, 60/92 for Emotional, 68/100 for Social, 64/96 for Work, 56.7/93.3 for Psychosocial, and 56.5/92.4 for Total. The Social scale demonstrated a marked ceiling effect where the 85th percentile score corresponded to the maximum, while an upper-end compression without a formal ceiling was observed for the Physical and Work scales. The distributions of the Emotional, Psychosocial, and Total scales, on the other hand, were broader with heavier lower tails.

**Figure 1:**
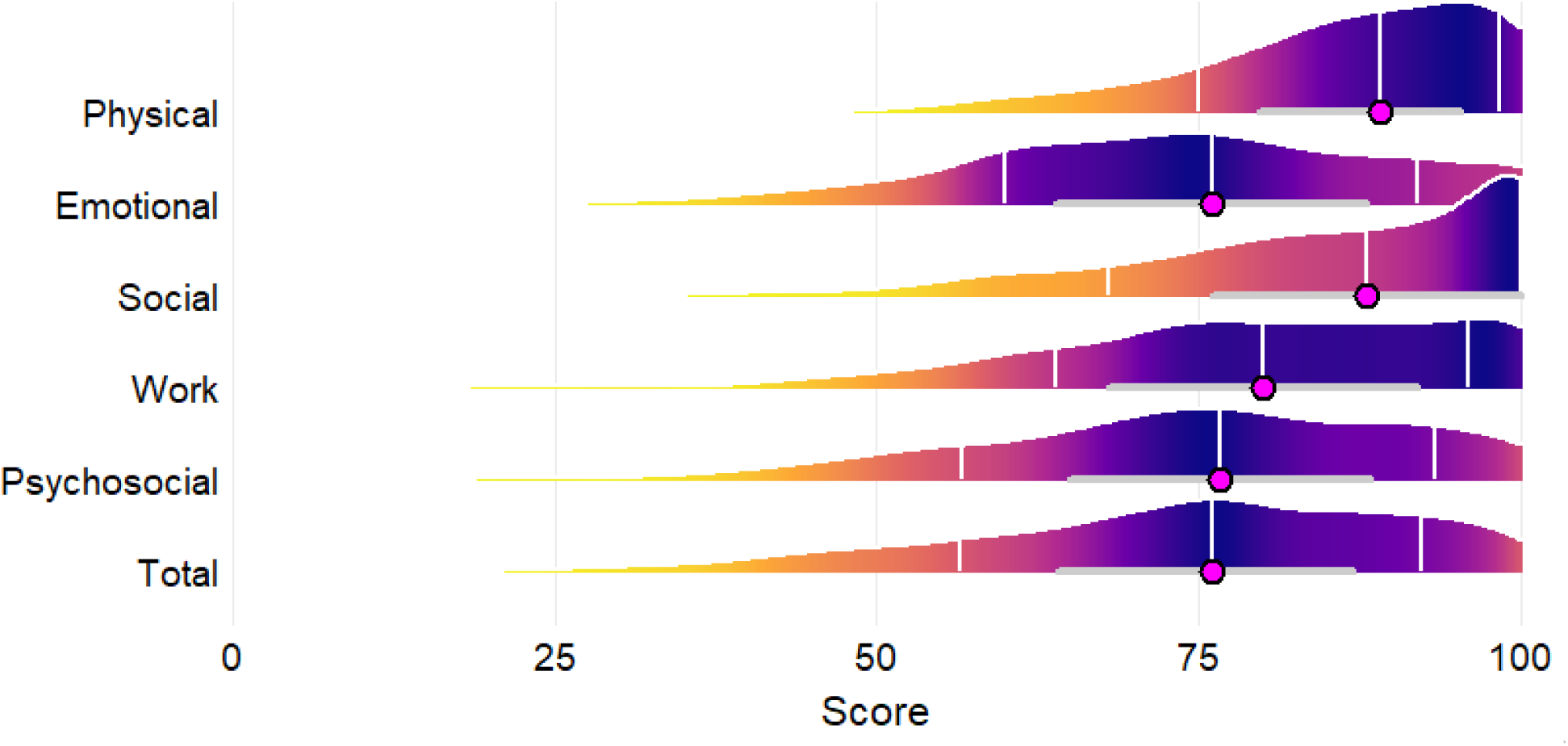
Score distributions for PedsQL Core scales (n = 868). Half-violin ridgelines display densities of scale scores (0–100; higher = better QoL), with the color gradient indicating relative density (yellow = low; purple = high). Vertical lines (white) mark the 15th, 50th (median), and 85th percentiles; horizontal bars (grey) show the interquartile range with a point (magenta) at the median. For each scale, the 15th and 85th percentiles respectively define the thresholds of its lower and upper bands.

#### Stage I: Representation of CHD complexity across HRQL bands

CHD complexity was unevenly distributed across the lower (bottom 15%) and upper (top 15%) HRQL score bands within each PedsQL GCS scale (Table 2). The mild CHD group were consistently over-represented in the upper band and under-represented in the lower band, whereas the severe CHD group showed the opposite pattern. Under-representation in both lower and upper bands of the Total scale was observed for the severe CHD group only. Complete contingency tables and residual analyses are presented in Supplemental Material Table S1.

**Table 2:** Representation of mild CHD and severe CHD in the lower (bottom 15%) and upper (top 15%) HRQL bands for the PedsQL GCS.

| Scale | Lower band*<br>(bottom 15%) | Upper band*<br>(top 15%) | ES <sup>†</sup> [95% CI] | p-value |
| --- | --- | --- | --- | --- |
| Physical | Mild ↓, Severe ↑ | Mild ↑, Severe ↓ | 0.10 [0.06, 0.15] | <0.001 |
| Emotional | Mild ↓, Severe ↑ | Mild ↑, Severe ↓ | 0.06 [0.00, 0.12] | 0.051 |
| Social | Mild ↓, Severe ↑ | Mild ↑, Severe ↓ | 0.09 [0.05, 0.14] | 0.001 |
| Work | Mild ↓, Severe ↑ | Mild ↑, Severe ↓ | 0.05 [0.00, 0.11] | 0.095 |
| Psychosocial | Mild ↓, Severe ↑ | Mild ↑, Severe ↓ | 0.09 [0.03, 0.14] | 0.002 |
| Total | Mild ↓, Severe ↓ | Mild ↑, Severe ↓ | 0.10 [0.05, 0.15] | <0.001 |

\* Arrows show deviation from expected counts for that CHD complexity: ↑ over-represented, ↓ under-represented. Full cell counts and standardized residuals are in Supplementary Table S1.
<sup>†</sup> Effect size estimate (Cramér's V).

Effect size estimates indicated small yet consistent associations. Cramér’s V ranged from 0.05–0.10 across all scales, with CIs excluding zero for the Physical (0.10 [0.06–0.15]), Social (0.09 [0.05–0.14]), Psychosocial (0.09 [0.03–0.14]), and Total (0.10 [0.05–0.15]) scales. The Emotional (0.06 [0.00–0.12]) and Work (0.05 [0.00–0.11]) scales however showed the same direction of effect but with wider uncertainty; their CIs including zero. Although the magnitudes of effect were modest, consistency across the scales indicates that more complex CHD is associated with a greater likelihood of worse HRQL.

#### Stage II: Predictors of overall HRQL at the lower and upper bands

For the Total Scale, QR modelling estimated how the prespecified predictors related to distributional effects on HRQL at the conditional 15th and 85th percentiles (Figure 2). Detailed estimates for all predictors are presented in Supplemental Material Table S2.

**Figure 2:**
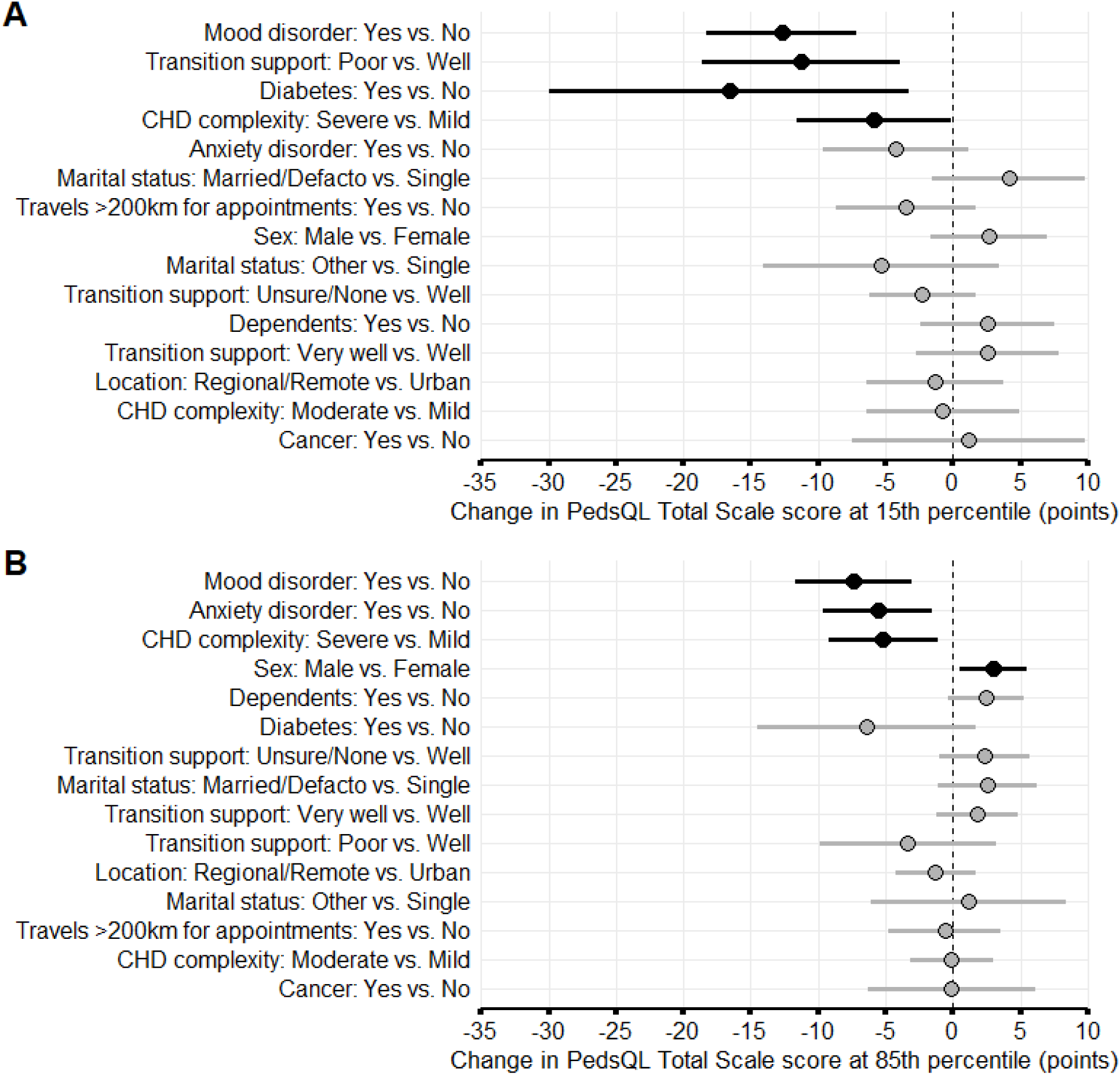
Quantile regression estimates of associations between predictors and PedsQL Total Scale scores (Stage II) at the conditional 15^th^ (A) and 85th percentiles (B). Points are coefficient estimates with 95% CIs. Black points/bars denote estimates whose 95% CIs do not cross 0 (statistically significant at α=0.05); grey points/bars denote estimates whose 95% CIs do cross 0. Positive values reflect an upward shift in the percentile score (better HRQL), and negative values reflect a downward shift (worse HRQL).

At the 15th percentile (lower band), several predictors were associated with worse HRQL. Patients with a mood disorder scored 12.7 points lower (95% CI −18.3 to −7.1), those reporting “poor” transition support scored 11.2 points lower (95% CI −18.6 to −3.9), and those with diabetes scored 16.6 points lower (95% CI −29.9 to −3.3). Severe CHD was also linked to lower scores (−5.9 points; 95% CI −11.6 to −0.1). Estimates for the remaining predictors (anxiety disorder, marital status, dependents, travel distance, location, and cancer history) were small or imprecise, with CIs including zero.

At the 85th percentile (upper band), a different picture emerged. Severe CHD again related to worse HRQL (-5.2 points; 95% CI -9.2 to -1.1), and mood disorder (-7.3 points; 95% CI - 11.7 to -3.0) and anxiety disorder (-5.6 points; 95% CI -9.7 to -1.5). In contrast, male sex was associated with better HRQL (higher scores) (+3.0 points; 95% CI 0.5 to 5.5). Other predictors at the upper tail (dependents, diabetes, transition support categories, location, marital status, travel distance, and cancer history) again showed small and statistically uncertain effects.

Across both tails, one age-spline term reached nominal significance at the 85th percentile; however, joint Wald tests of all spline terms found no evidence of a nonlinear age effect, so age was not retained as a predictor at either tail. Model adequacy and linearity assumptions were satisfied, and sensitivity analyses yielded consistent results.

#### Stage III: Predictors of overall HRQL upper-band membership within the severe CHD group

For the severe CHD group (n=218), Bayesian PO modelling showed several sociodemographic and clinical predictors of HRQL upper-band membership in the Total Scale (Figure 3). Therein, the ORs for each predictor are shown in panel A, and their corresponding posterior probabilities in panel B. In the following delineation, we denote “Pr(upper-band)” as the posterior probability of membership in the HRQL upper-band for a predictor; values ≥0.95 indicate strong evidence of higher probability, values ≤0.05 indicate strong evidence of lower probability, and values ≈0.5 indicate no clear directional evidence. In this model, OR > 1 indicates better HRQL (shift towards the upper-band) and OR < 1 indicates worse HRQL (shift towards the middle- and lower-bands). Model diagnostics supported proportional-odds assumptions, and convergence and fit criteria were satisfactory.

**Figure 3:**
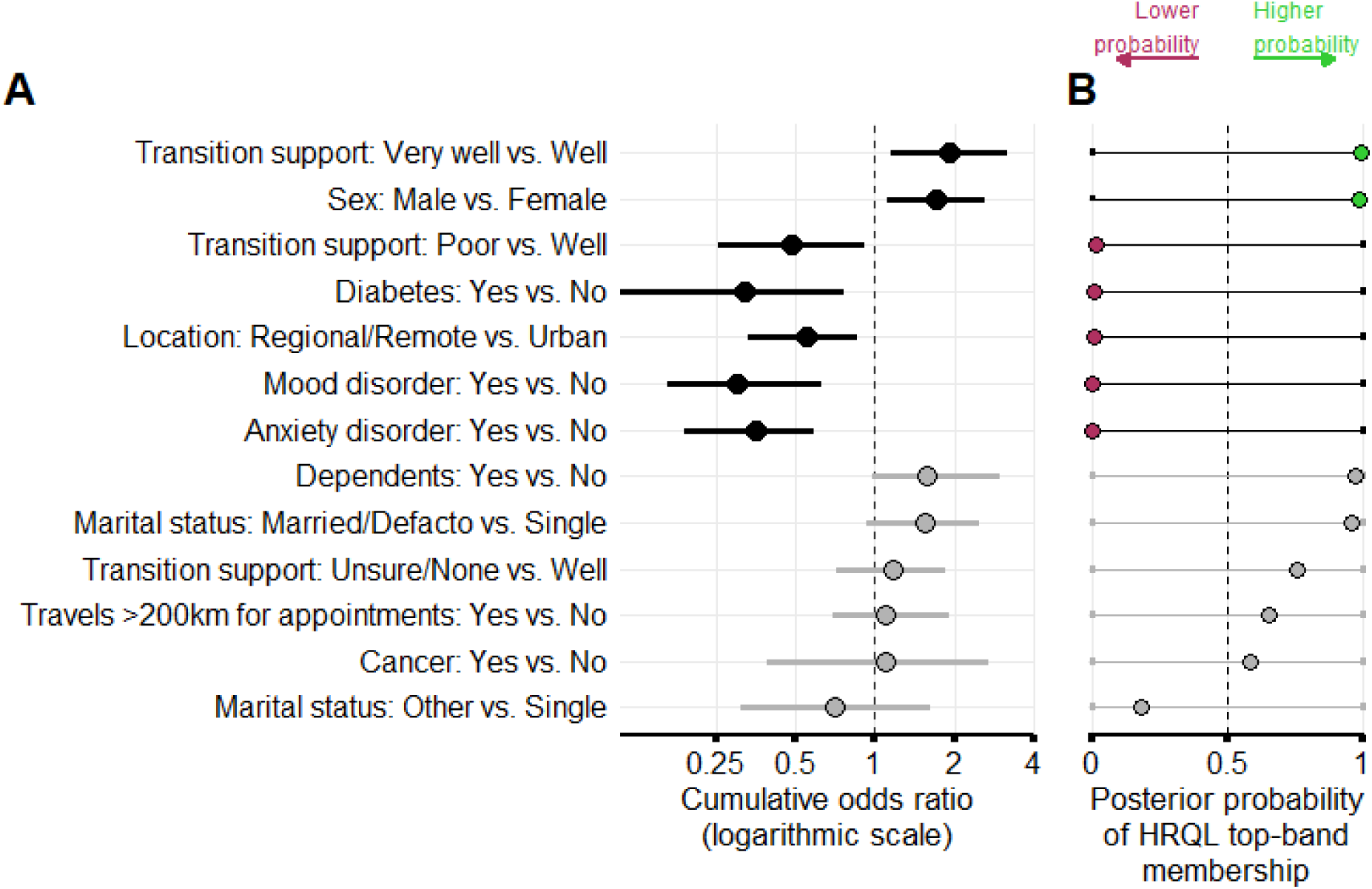
Predictors of HRQL band membership within the severe CHD subgroup (Stage III). **Panel A:** Posterior medians of cumulative odds ratios with 95% credible intervals on a logarithmic scale; values >1 indicate greater odds of being in the HRQL upper-band, and values <1 indicate greater odds of being in the HRQL lower-band. Black points/lines denote estimates whose 95% credible intervals do not include 1; grey denote intervals that do. **Panel B:** Posterior probability that each predictor raises the chance of HRQL upper-band membership. The dashed vertical line at 0.5 marks no directional evidence. Points to the right of 0.5 (green) indicate the predictor is more likely to increase upper-band membership; points to the left (maroon) indicate it is more likely to decrease it.

Patients who reported they were “very well” supported during transition had markedly better HRQL. Their odds of better overall HRQL were nearly doubled (OR = 1.91; 95% CrI 1.15 to 3.18), and the posterior probability that this predictor increases the chance of upper-band membership was essentially certain (Pr(upper-band) = 0.99). In practical terms, “very well supported” patients are far more likely to sit in the best HRQL band than otherwise similar peers.

A similar pattern was seen for male sex: the association favored better HRQL (OR = 1.71; 95% CrI 1.11 to 2.59) with a correspondingly high probability of increasing upper-band membership (Pr(upper-band) = 0.99). Thus, for sex, both the direction (panel A) and the strength of evidence (panel B) align.

In contrast, poor transition support pulled outcomes the other way. The odds ratio indicated worse HRQL (OR = 0.48; 95% CrI 0.25 to 0.92), and the probability of increasing upper-band membership was very low (Pr(upper-band) = 0.06), reinforcing that poor support is acts as a barrier to achieving higher HRQL strata.

Several clinical and contextual factors also showed consistent evidence of worse HRQL: diabetes (OR = 0.32; 95% CrI 0.11 to 0.77; Pr(upper-band) = 0.01), regional/remote location (OR = 0.55; 95% CrI 0.33 to 0.86; Pr(upper-band) = 0.01), mood disorder (OR = 0.30; 95% CrI 0.16 to 0.63; Pr(upper-band) = 0.01), and anxiety disorder (OR = 0.36; 95% CrI 0.19 to 0.59; Pr(upper-band) = 0.00). For each of these predictors, panel A shows odds ratios below 1 (shifts toward the middle- and lower-HRQL bands), and panel B shows posterior probabilities close to zero, indicating strong evidence against increasing upper-band membership.

The remaining predictors (dependents, marital status, travel distance, and cancer history) had estimates that were directionally consistent but statistically uncertain–their CrIs included 1 and Pr(upper-band) values hovered near 0.5, indicating no clear directional evidence.

Together, the two panels convey both effect size/direction (panel A) and the strength of evidence for increasing HRQL upper-band membership (panel B), highlighting transition support, sex, diabetes, mental health, and location as the most informative predictors in this severe CHD cohort. Detailed estimates can be found in Supplemental Material Table S3 where we also present average marginal effects (AMEs) as absolute percentage-point (pp) changes in the probability of being in the lower or upper bands for each predictor variable.

## Discussion

### Major novel findings

In a large cohort of Australian adults with CHD, self-reported HRQL was generally high but unevenly distributed across domains, with clear ceiling effects in Social and Physical functioning and longer lower tails for Emotional, Psychosocial, and Total Scale scores. Across those distributions, CHD complexity showed the expected gradient: severe lesions were over-represented among patients with worse HRQL and under-represented among those with best HRQL.

Utilizing the PedsQL Total Scale as a measure of overall HRQL, QR modelling revealed severe CHD and mood disorder to be consistently linked with worse outcomes, implicating anatomical complexity and depression as cross-spectrum risks. Among patients with the lowest scores (lower tail), poor transition support and diabetes had the greatest additional impact, highlighting potential areas for intervention. Even among patients with higher scores (upper tail), anxiety was associated with lower HRQL, while men reported slightly better outcomes. Moderate CHD, age, and most other sociodemographic factors showed little or uncertain influence after adjustment.

Within the severe CHD subgroup, Bayesian PO modelling highlighted clear and actionable associations. Two predictors aligned consistently with better overall HRQL: male sex and “very well”-supported transition from pediatric to adult cardiovascular care; each carry a high probability of increasing the chance of upper-band membership. In contrast, poor transition support, mood or anxiety disorder, diabetes, and regional/remote location all showed low probability of improving (and strong probability of worsening) upper-band membership, indicating reliably detrimental associations beyond anatomic severity. Clinically, these findings argue for structured transition pathways, integrated mental-health care, targeted diabetes management, and strengthened access for regional/remote patients as priorities to improve the *overall* quality of survival in adults with severe CHD.

### Context compared with prior work

The demography of CHD continues to shift toward adulthood as survival improves, with adults now comprising roughly two-thirds of the total CHD population and adult prevalence rising markedly in the 2000–2010 decade^5^. These population changes, coupled with accumulating comorbidity, emphasize the need to move beyond mortality and procedure counts to PROs such as HRQL.

Prior Australian work in pediatric cohorts linked greater anatomic/physiologic severity to worse HRQL, especially in physical and psychosocial domains, and highlighted the broader family impact of complex disease^10^. Our findings extend this gradient into adulthood but also reveal important heterogeneity: despite generally high overall HRQL scores, a distinct subgroup experiences very low emotional and psychosocial functioning, indicating that anatomic severity alone is not sufficient to explain lived outcomes in adult CHD. Earlier international studies showed that PROs vary by New York Heart Association functional class and socioeconomic context and that health-system characteristics can shape reported HRQL across countries^11,12^.

Our study advances this literature on three fronts. First, we analyze a large, registry-linked national ACHD cohort, improving precision compared to single-center series. Second, we move decisively beyond mean-based comparisons by applying quantile regression to characterize how predictors shape the lower and upper tails of the overall HRQL distribution, and by using a Bayesian proportional-odds model to quantify probabilities of upper-band membership. To our knowledge, this pairing with the PedsQL Generic Core Scales in ACHD is among the first of its kind, allowing us to detect predictors that selectively worsen or improve outcomes; patterns that conventional models often miss. The Bayesian framework further strengthens clinical interpretation in cohort sizes typical of ACHD research by providing transparent probabilities of benefit or harm rather than relying solely on arbitrary significance thresholds; this yields decision-ready, directionally clear evidence even when classical power is limited. Third, the Australian setting is uniquely informative: national Medicare coverage attenuates socioeconomic gradients in healthcare access, allowing observed predictors such as transition experience, mental-health comorbidity, regional/remote location, and diabetes to be interpreted as ACHD-specific targets rather than artifacts of unequal utilization.

Together, these features position our work as a distribution-aware, probability-based, and health-system-contextualized contribution that identifies actionable levers to improve the quality of survival in adult CHD.

### Interpretation and clinical implications

First, distributional heterogeneity matters. Mean HRQL can appear reassuring while a clinically important minority experiences very low scores. By targeting the tails with quantile regression and confirming probability of upper-band membership with the Bayesian model, we identified who is at risk and which predictors co-travel with worse HRQL.

Second, transition quality is not merely an organizational metric: patients who perceived poor support during transfer from pediatric to adult care had markedly lower HRQL, and those living in regional/remote areas were additionally disadvantaged. These findings support investment in structured transition pathways, outreach, and telehealth capacities for geographically dispersed populations.

Third, mental-health comorbidity is a high-priority target. Mood and anxiety disorders were consistently and statistically significant predictors of worse HRQL and associated with lower scores at both tails and a high probability of reducing upper-band membership. Routine screening, rapid referral pathways, and integrated psychological care should be considered core components of ACHD services rather than adjuncts.

### Strengths and limitations

Strengths of this study include a large, registry-linked national cohort; prespecified predictors spanning clinical, psychosocial, and access domains; and complementary modelling that interrogates both continuous distributional tails (quantile regression) and ordered band membership (proportional-odds). Our focus on extremes improves clinical relevance by identifying patients most likely to need support.

Limitations include the cross-sectional design, which precludes causal inference, and potential residual confounding. Although we adjusted for anatomy/physiology via CHD complexity strata, we did not model granular lesion-specific features, interventions, or hemodynamic metrics that may further stratify risk. Finally, while international studies have highlighted health-system effects on PROs, our single-country design may conversely be advantageous. Medicare coverage in Australia attenuates disparities in healthcare access across socioeconomic classes, strengthening the inference that the predictors identified in this study are more reflective of ACHD-specific challenges rather than inequities in healthcare utilization.

### Future directions

Prospective studies should investigate whether improving transition processes, expanding regional/remote access, and integrating mental-health assessment and care improve HRQL, particularly for patients with the lowest scores. Linking PROs with clinical trajectories (arrhythmia burden, reinterventions, pregnancy outcomes, and employment/education) could clarify mechanisms and help define meaningful change thresholds for ACHD programs. Cross-jurisdictional comparisons using harmonized methods would also help disentangle patient-level from system-level drivers of better or worse HRQL.

## Conclusions

In contemporary Australian adults with CHD, overall HRQL is generally high, yet a substantial subgroup experiences markedly poor outcomes. By applying distributional methods, quantile regression and Bayesian proportional-odds modelling, our study is the first to move beyond mean differences to identify predictors that contribute to better or worse HRQL in the ACHD population. The predictors we found extend beyond anatomy to include transition experience, mental-health comorbidity, diabetes, and regional/remote residence, while the context of Australian Medicare coverage reduces socioeconomic confounding. These findings highlight that the predictors driving very low HRQL are modifiable and directly actionable. Embedding structured transition pathways, routine mental-health screening and treatment, and strategies to improve access for regional/remote patients should be regarded as core components of ACHD care. As the ACHD population continues to grow, improving the quality of survival requires targeted, system-level interventions that complement surgical and medical successes.

## Data Availability

The authors will not make study data openly available. Any future use of de-identified data will require specific ethics and governance approvals and will be limited to approved collaborators.

## Acknowledgments

This study used data from the National Australian Congenital Heart Disease Registry, with support from HeartKids Australia, to assess the health-related quality of life of adults living with CHD in Australia. The Registry is a collaborative initiative developed with guidance from the Congenital Heart Alliance of Australia and New Zealand, whose clinicians and researchers contributed expertise to its establishment and to the design of this study. HeartKids Australia, the leading consumer body for childhood heart disease, continues to play an integral role by ensuring patient perspectives are represented and by advocating for future research.

## Supplemental Material

Figure S1

Tables S1–S3

